# The interplay between pre-season population susceptibility and the effectiveness of the rolled out vaccination control the timing and size of an emerging seasonal influenza wave in England

**DOI:** 10.1101/2023.01.06.23284264

**Authors:** E. van Leeuwen, J. Panovska-Griffiths, S. Elgohari, A. Charlett, C. Watson

## Abstract

Relaxing social distancing measures and reduced level of influenza over the last two seasons may lead to a winter 2022 influenza wave in England. We used an established model for influenza transmission and vaccination to evaluate the rolled out influenza immunisation programme over October to December 2022. Specifically, we explored how the interplay between pre-season population susceptibility and influenza vaccine efficacy control the timing and the size of a possible winter influenza wave. Our findings suggest that susceptibility affects the timing and the height of a potential influenza wave, with higher susceptibility leading to an earlier and larger influenza wave while vaccine efficacy controls the size of the peak of the influenza wave. With pre-season susceptibility higher than pre-COVID-19 levels, under the planned vaccine programme an early influenza epidemic wave is possible, its size dependent on vaccine effectiveness against the circulating strain. If pre-season susceptibility is low and similar to pre-COVID levels, the planned influenza vaccine programme with an effective vaccine could largely suppress a winter 2022 influenza outbreak in England.

## 2. Background

Since early 2020, alongside large-scale vaccination against COVID-19, social distancing measures imposed by governments have been widely used to mitigate the transmission of SARS-CoV-2 and its variants. The reduction of social mixing have also reduced the level of other infectious diseases such as influenza, with reported low levels of influenza worldwide over the 2020/21 and the 2021/22 seasons [4, 5, 16]. Although, influenza subtypes skipping seasons is not uncommon, having two consecutive influenza seasons with very low incidence is rare [4]. While influenza is a common infection and mild for most people, it can be very dangerous for vulnerable people, including older adults, pregnant women and people with underlying health conditions, for whom infection can result in hospitalisation and death. Seasonal influenza can also put winter pressure on the National Health Service (NHS), the extent of which depends on a number of factors including whether there are other circulating viruses such as SARS-CoV-2 and respiratory syncytial virus (RSV) which can induce increased hospitalisations, the state of the backlog of elective surgeries following the pandemic years, increased demand for GP consultations and hospitalisations and intensive care bed admissions. Even in absence of other high prevalence circulating viruses, influenza can put constrains on the NHS; in the winter 2017–2018, influenza levels were high and this led to deferral of all elective inpatient and outpatient NHS care in England throughout January 2018 [6],[8].

Social distancing measures relaxing, either fully or partly, and possible increased susceptibility to influenza viruses this year, may lead to an increased influenza season in 2022. Australia’s season is often examined as a portent for northern hemisphere activity. Over April-July 2022, Australia experienced an early influenza season. This is likely to reflect factors such as national COVID-19 control circumstances and influenza vaccination coverage, but indicates the potential for disrupted seasonality.

### 2.1. Planned vaccination strategy for 2022/23 season in England

Vaccination against the circulating influenza strain is the most effective way to prevent influenza surge in the population, and England has a well-established annual influenza immunisation programme rolled out from September each year. This annual influenza immunisation programme is delivered to protect people at risk from influenza and is the most important public health intervention to mitigate influenza resurgence, reduce morbidity and mortality and winter pressure on the NHS. Immunisation is needed annually because of waning protection and the virus evolves with different viral strains circulating in the population each year.

Eligible cohorts and vaccine types are decided based on scientific advice from the Joint Committee on Vaccination and Immunisation (JCVI). In 2022/23, the agreed cohorts were children age 2 or 3 year old on 31 August 2022, primary school aged children (4-11 years old) and secondary school children to Year 9 (11-13 years old) with some older children also included, those aged 6 months to under 65 years in clinical risk groups, and those aged 50 years and over. This is an increase in eligible cohorts compared to the pre-pandemic (2019/20) influenza immunisation programme, which did not include healthy children 11-13 years old nor healthy adults between 50 and 64. A(H3N2) subtype is currently detected more frequently than A(H1N1)pdm09 in sentinel laboratories in England. Influenza vaccine strains are updated to better match circulating strains. Virological surveillance shows the updated A(H3N2) 2022-23 vaccine component to be well matched to the circulating strain.

Here, we evaluate whether this planned vaccination strategy will be able to mitigate an influenza epidemic wave over the autumn and winter 2022 in England, using an established epidemiological model for influenza transmission and vaccination [2, 13, 14]. Our analyses have provided informed modelling advice to the Scientific Pandemic Influenza Group on Modelling sub-group (SPI-M) and the Scientific Advisory Group for Emergencies (SAGE) in September 2022 on the possible winter pressures in the UK.

## 3. Methods

We quantify the timing and the size of an influenza epidemic wave peak under different susceptibility and vaccine efficacy scenarios. Specifically, we simulate two scenarios of pre-season population susceptibility (low and similar to pre-COVID levels versus high due to non-exposure to/low prevalence of influenza over last two years) and two scenarios of vaccine efficacy (low and high; with details in Figure 2A). We also assume vaccine uptake to be based on the 2021/22 uptake [11, 12] (Figure 2B).

### 3.1. Transmission model

We are using an established epidemiological model for influenza transmission and vaccination [2, 13, 9]. The model has been re-fitted to data for the 2017/18, following [9], but with updated vaccine effectiveness parameters [7]. Using the posterior parameter values, we can model the impact of the updated vaccination programme and assuming different scenarios for vaccine effectiveness and population susceptibility.

### 3.2. Hospitalisation model

The hospitalisation model assumes that infections result in hospitalisations with an age group dependent rate *α*_*i*_. We also assume that there could be a delay between being infected and hospitalisations (*γ*). This is modelled by including two delay states in the compartmental model (Δ*i, j*), resulting in gamma distributed delay times:

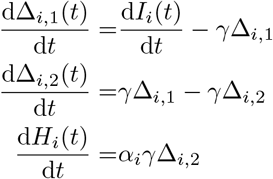

The data we have are the weekly number of hospitalisations (*k*_*i,t*_) by age group and the size of the monitored population (*n*_*i,t*_). The latter can change dependent on the number of hospitals that report their cases by week. The likelihood function is then defined as follows. Note that we calculate the modelled number of hospitalisation by integrating over the previous week, resulting in the total number of hospitalisations for that week.

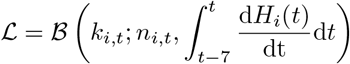

Our original infection model results in posterior samples for the weekly number of infections. For the hospitalisation model we therefore assume that during that week the number of new infections is constant over time. Inference of the parameters for the hospitalisations model is done by fitting the above model separately to a subsample of the posterior samples using MCMC. This results in a number of posterior samples for the hospitalisations model, of which we store one (i.e. each posterior sample of the infection model results in a corresponding sample for the hospitalisations model). While this approach does not result in a proper posterior sample, because it does not account for the fact that some of the posterior samples of the infection model are more likely given the hospitalisation data, it should be a conservative approximation of the posterior parameters given the hospitalisation data.

### 3.3. Serological data

We have limited serological data available, making it difficult to compare the two seasons. For example, the 2017 data was based on opportunistic sera samples, while the 2021 data is based on samples collected through the RCGP [7, 15]. For the 2017 data we did not have their vaccination status available, while for the 2021 season we do not have data for the younger age groups. To get an indication of the difference in susceptibility, we looked at the total percentage of samples with a titre of 40 or over for the most relevant strains. For 2017 we used the HongKong_TC strain, while for current values we relied on the Bangladesh strain results. Overall, this gave us a relative susceptibility of 1.1 for the high susceptibility scenario. Due to lack of subtype B specific data, we will be assuming the same value for subtype B.

## 4. Results

### 4.1. The influenza burden over the pandemic years

During the 2020-2022 pandemic years, influenza levels in England declined dramatically (Figure 1). We show that the peak rate of influenza hospitalisations was 5.3 and 6.9 times lower in 2021/22 compared to 2018/19 and 2017/18 respectively and close to zero in 2020/21 (Figure 1). At the time of writing this, in November 2022, the growth has been consistent for a number of weeks, with a slightly higher hospitalisation rate than in previous seasons [10]. Still, we have not yet seen the rapid increase in cases that we normally see at the start of the influenza wave. Instead, the increase in cases has been relatively steady [10].

**Figure 1:**
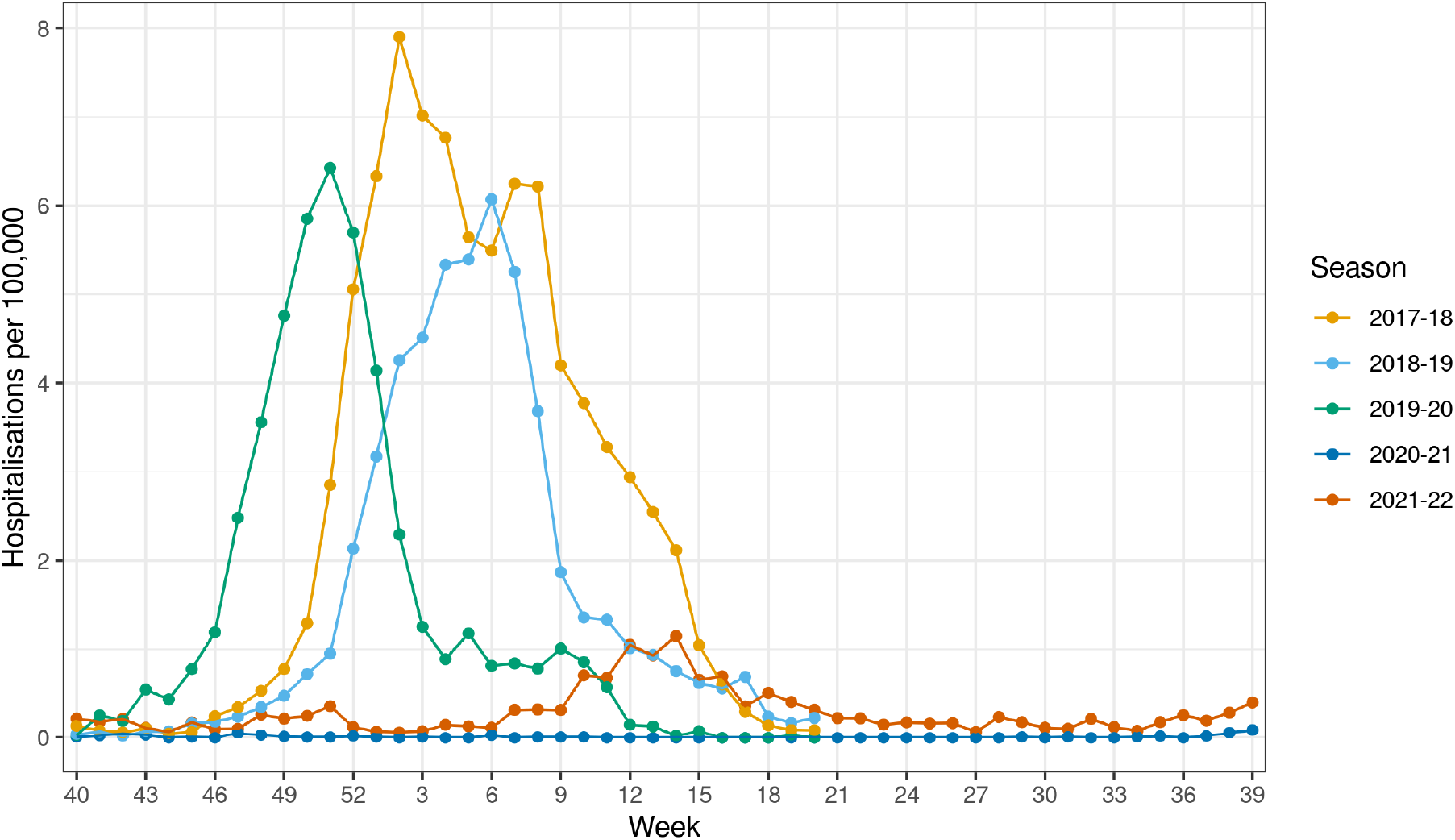
Historic rate of influenza hospitalisation (all levels of care) by week of admission and season, all ages, using sentinel data from acute NHS trusts in England. The rates are based on the catchment population of responding trusts in the sentinel surveillance scheme. In 2017/18 both AH3N2 and B were circulating in significant numbers, 2018-19 was dominated by AH1N1pdm09, while 2019-20 was again dominated by AH3N2.

### 4.2. Emerging timing and size of winter influenza waves depends on population susceptibility and vaccine efficacy

Using an established epidemiological model for influenza transmission and vaccination combined with data for England[1], we explored how the interplay between population susceptibility and influenza vaccine efficacy impact the timing and the size of a possible winter influenza wave over the 2022/2023 season.

Our results suggest that the combination of relaxing the COVID-19 social distancing measures and two previous low incidence influenza seasons in England could lead to a large influenza epidemic wave in late 2022 (Figures 3). The size and the timing of such an influenza epidemic wave in late 2022 in England are highly dependent on the population susceptibility, vaccine uptake and vaccine efficacy (Figure 2A-C).

**Figure 2:**
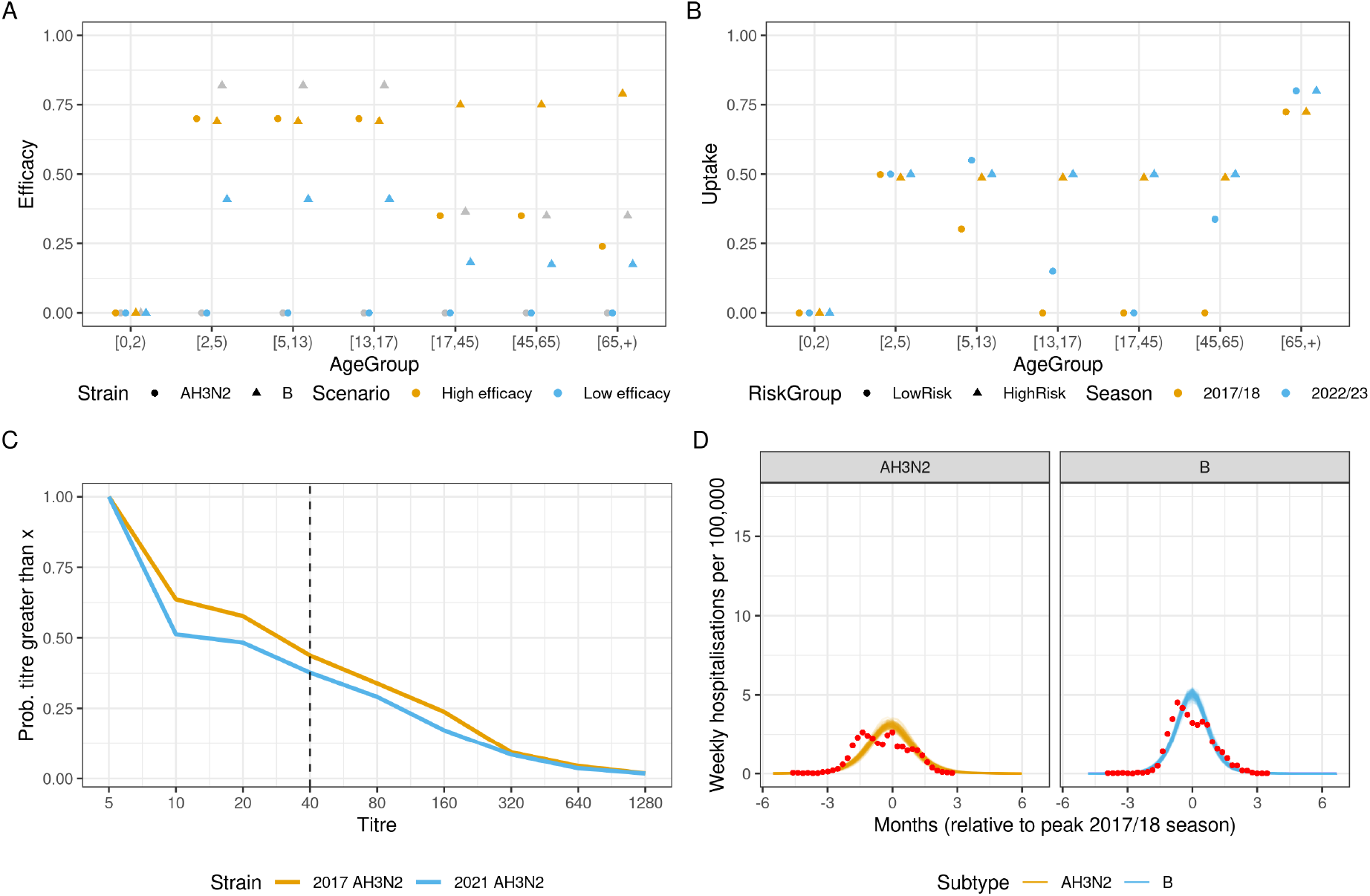
Vaccination assumptions used in the scenarios. A) High efficacy assumptions for AH3N2 are based on 2021/22 results, while for subtype B we use the results from a meta-analysis [3]. Low values are half the estimated efficacy in the 2017/18 season (shown in grey for reference; [7]. B) Vaccine uptake is based on 2021/22 uptake, which includes uptake in the additional eligible cohorts. C) Available serological data at the start of both seasons. The lines represent the probability that a sample has the given titre value (x-axis) or higher. Higher values correspond with a higher proportion of the population having antibodies against influenza. The dashed line shows the titre of 40. Generally, it is assumed that 50 percent of individuals with a titre value of 40 or above are immune. D) The fitted hospitalisation rates for both H3N2 and B. The red points show the data in 2017/18.

**Figure 3:**
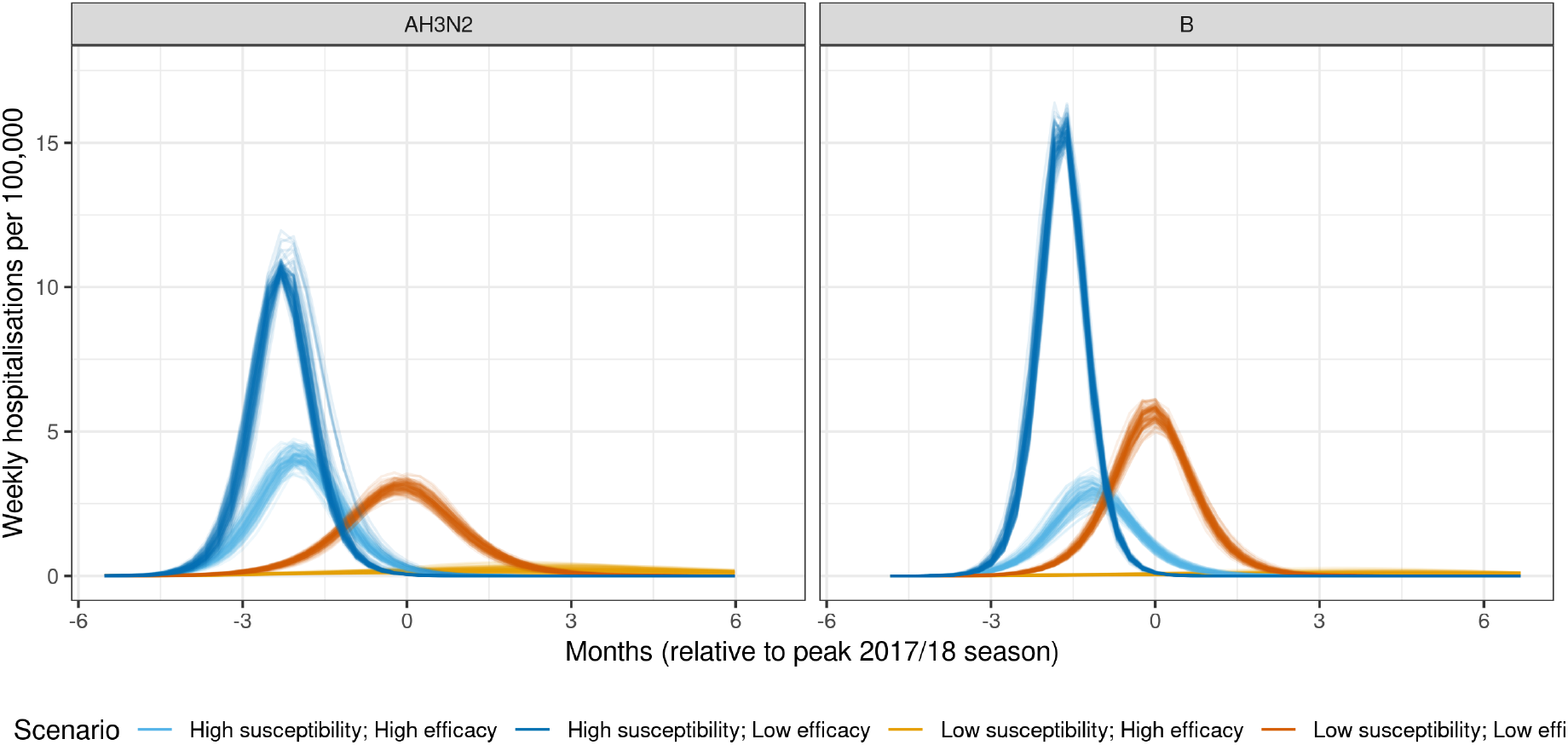
Inferred weekly hospitalisations by age group per 100,000 individuals. The colour labels refer respectively to the different scenarios. All the scenarios assume the projected vaccine uptake, with either, high susceptibility (blue) or low susceptibility (orange). Vaccine efficacy is assumed to be low (lighter colours) or high. Each trace represents a simulation (total of a 100 simulations per scenario).

Population susceptibility affects the timing and the size of a potential influenza wave, with higher susceptibility leading to an earlier influenza wave (comparing the two blue curves with the two orange curves in Figures 3 and 2D). The efficacy of the rolled out vaccine affects the magnitude of the peak of the influenza wave, with more effective vaccine able to flatten the epidemic curve substantially (comparing the two blue curves with each other and the two orange curves with each other in Figure 3 and comparing scenarios in Table 1).

**Table 1.** Peak hospitalisations per 100,000 in the different scenarios as well as the relative timing of the peak timing (in months) compared to the base case (2017/18) season. Negative values indicate that the peak is projected to fall before the 2017/18 peak.

| Subtype | Susceptibility | Efficacy | Peak | Peak time |
| --- | --- | --- | --- | --- |
| AH3N2 | High | High | 4.134 [3.48,4.744] | -2.07 [-2.3,-1.61] |
|  |  | Low | 10.621 [10.161,11.963] | -2.3 [-2.3,-2.07] |
|  | Low | High | 0.198 [0.075,0.491] | 2.3 [-1.61,3.45] |
|  |  | Low | 3.115 [2.69,3.575] | 0 [-0.23,0] |
| B | High | High | 2.932 [2.217,3.745] | -1.15 [-1.38,-0.92] |
|  |  | Low | 15.423 [14.698,16.385] | -1.61 [-1.84,-1.61] |
|  | Low | High | 0.074 [0.025,0.278] | 3.68 [-1.61,5.29] |
|  |  | Low | 5.709 [4.969,6.133] | 0 [-0.23,0] |

We show that if the population susceptibility is similar to that in 2017/18 season i.e. pre-COVID-19, the extended planned influenza immunisation programme for 2022/23 with an effective vaccine has the potential to prevent an influenza resurgence over September-December 2022 (light orange curve in Figure 3 and Table 1). Increased susceptibility can lead to an earlier influenza wave (blue shaded curves in Figure 3 also see Table 1) while vaccine efficacy controls the size of the peak of the influenza wave (light shaded peaks in Figure 3)

## 5. Discussion

We use an established model for transmission and vaccination against influenza in England, to evaluate whether the rolled out influenza immunisation strategy can prevent a large epidemic wave over the 2022/2023 season. This work was presented and discussed at a meeting by SPI-M in September 2022 the UK. Pre-COVID-19 i.e pre the 2020/2021 influenza season, our model has been routinely used to explore the impact of different mixing patterns and the planned immunisation programme on the yearly upcoming influenza wave, with our results shared with the scientific advisory groups SPI-M and SAGE in the UK.

Aiming to evaluate the impact of the planned influenza vaccine in England, we explore how the interplay between pre-season population susceptibility and influenza vaccine efficacy affect the timing and the size of a possible winter influenza wave in England over October-December 2022. Our findings suggest that susceptibility affects the timing and the height of a potential influenza wave, with higher susceptibility leading to an earlier and higher influenza wave while vaccine efficacy controls the size of the peak of the influenza wave. We observed that influenza activity substantially declined during the 2020–21 and the 2021/2022 seasons and during the COVID-19 pandemic in England. As a consequence, our findings highlight that an extensive annual influenza immunisation programme can substantially mitigate the impact of an influenza epidemic in a season where the underlying population immunity is reduced.

At the time of writing this, November 2022, in England, there have been indications of an early onset of the 2022/2023 influenza season, with the hospital admission rate for confirmed cases of influenza in mid-October being greater in 2022 than seen in preceding years within norms for influenza seasonality [10]. Even with this in mind, our findings suggest that looking ahead there is uncertainty as to the size of the upcoming winter influenza epidemic wave over the 2022/2023 season. The importance and novelty of our work is that we show, under different scenarios, a spectrum of possible winter pressures scenarios from influenza over this season: from a very small influenza epidemic wave with highly effective vaccines and low population susceptibility to a very large epidemic wave with low effectiveness vaccines and high population susceptibility. This uncertainty has also been shown in other modelling studies. For example, analysis by Ali et al. [1] suggests that there is a possibility of large upcoming influenza seasons worldwide, while Felix Garza et al. [4] suggest that future seasonal influenza virus epidemics may likely be similar to the previous one and would not have an increased burden. Such a broad spectrum of results and uncertainity is an important aspect of modelling and often a consequence of different modelling assumptions and methods used.

The uniqueness of our study is that we show that the uncertainty of the timing and height of a possible influenza epidemic wave can emerge from the interplay between pre-season population susceptibility and the rolled-out vaccine effectiveness. Bearing in mind that we don’t know the vaccine effectiveness for the season until the end of the influenza season, our modelling remains a crucial tool for pre-season winter pressure planning.

As with any modelling work, our study has some limitations. Firstly, we note that the modelling is based on the assumption that social contacts rates will return to pre-COVID-19 levels for this winter. Current data suggest that contact rates are still lower than they used to be pre-pandemic (especially in adults). If this continues throughout the season, then that could suppress influenza transmission further, especially in the adults age groups. Secondly, we note that we assume that the majority of the vaccine doses will be distributed before any influenza outbreak. In case of a very early peak, the planned immunisation programme could be much less effective, because it arrives after the start of the outbreak.

As we transition from the pandemic COVID-19 era, there is still uncertainty around changes to seasonal activity of many respiratory infections, including any timescales over which these might reestablish conventional seasonality. Further work is needed across surveillance and modelling, including accounting for any increases in microbiological testing.

Modelling using epidemiological and statistical models has been a crucial aspect of the informed advice for policy decision making over the COVID-19 epidemic in the UK. As we move to the “living with COVID-19 era”, expanding existing models, developing new ones and combining them with data remains an important tool for providing quantitative evidence for the outcome of possible interventions, including vaccination, to improve public health.

## 6. Conclusions

Using existing epidemiological model combined with influenza data from England, we determined that if susceptibility in the population is largely unchanged post COVID-19 and the rolled out vaccine is effective, then the planned vaccine programme could suppress any emerging influenza outbreaks. If susceptibility to influenza is higher than previous years, then an influenza epidemic is possible. Its timing depends on the susceptibility level, and its peak value depends on the effectiveness of the vaccine; this wave could be significantly worse than historic waves under cases of high susceptibility and poorly matches to the strain variant or could be negligible if susceptibility is the same as pre-COVID and the vaccine is well-matched to the circulating influenza strain.

## Data Availability

All data produced in the present study are available upon reasonable request to the authors.

## 7. Contribution

EvL, AC and CW conceived the study. EvL developed and undertook the modelling with input from JPG, AC, CW. The scenarios modelled were designed based on conversations within UK Health Security Agency (UKHSA). JPG and EvL wrote the manuscript, with input from AC, CW and SE. All authors approved the final version. EvL and JPG are the manuscript’s guarantors.

## 8. Acknowledgements

The authors would like to thank Dr. Katja Hoschler, the serology team of the Respiratory Virus Unit and Dr. Heather Whittaker for providing the data and guidance on analysing the serological data used to estimate susceptibility in this manuscript. EvL was supported by the National Institute for Health Research (NIHR) Health Protection Research Unit (HPRU) in Modelling and Health Economics, a partnership between UKHSA, Imperial College London, and LSHTM (grant number NIHR200908). JPG’s work is supported by the UK Health Security Agency and UK Department of Health and Social Care. The funders had no role in the study design, data analysis, data interpretation, or writing of the report. The views expressed in this article are those of the authors and not necessarily those of the UK Health Security Agency or the UK Department of Health and Social Care.

## Notes

### Competing Interest Statement

The authors have declared no competing interest.

